# A distributional regression approach to modeling the impact of structural and intermediary social determinants on communities burdened by tuberculosis in Eastern Amazonia – Brazil

**DOI:** 10.1101/2022.11.03.22281901

**Authors:** Clóvis Luciano Giacomet, Antônio Carlos Vieira Ramos, Heriederson Sávio Dias Moura, Thaís Zamboni Berra, Yan Mathias Alves, Felipe Mendes Delpino, Jason E. Farley, Nancy R. Reynolds, Jonas Bodini Alonso, Titilade Kehinde Ayandeyi Teibo, Ricardo Alexandre Arcêncio

**Affiliations:** Interunits PhD Program in Nursing, University of São Paulo College of Nursing at Ribeirão Preto, Ribeirão Preto, Brazil; Graduate Program in Public Health Nursing, University of São Paulo College of Nursing at Ribeirão Preto, Ribeirão Preto, Brazil; Postgraduate Program in Nursing, Federal University of Pelotas, Pelotas, Brazil; The Center for Infectious Disease and Nursing Innovation, Johns Hopkins School of Nursing, Baltimore, United States of America; Research Support Center, University of São Paulo College of Nursing at Ribeirão Preto, Ribeirão Preto, Brazil; Department of Maternal and Child Nursing and Public Health, University of São Paulo College of Nursing at Ribeirão Preto, Ribeirão Preto, Brazil

## Abstract

**Background:** TB is a disease affected by social determinants of health; however, it is unclear what its structural and intermediary determinants are in Eastern Amazonia. The region contains many natural resources, yet it suffers drastically from poverty, inequality, and neglected diseases. Here, we aimed to employ mathematical modeling to evaluate the influence of structural and intermediary determinants of health on TB in Eastern Amazonia – Brazil.

**Methods:** We conducted an ecological study. We considered cases diagnosed of TB and collected data by census tract to measure the social determinants. We applied the *generalized additive model for location, scale, and shape* (GAMLSS) framework to identify the effect of social determinants on communities with a high prevalence of TB. The Double Poisson distribution (DPO) was selected and we tested the inclusion of quadratic effects.

**Results:** 1,730 people were selected. The majority were female (59.3%), aged 31 to 59 years (47.6%), blacks (67.9%), schooling level of 5 to 8 years (18.7%). Prevalence of alcoholism was 8.6% and mental illness 0.7%. The GAMLSS analyses showed that the risk of community incidence of TB is associated with the proportion of the population without basic sanitation and also with the age groups 16-31 years and > 61 years.

**Conclusions:** The study revealed that GAMLSS is an strategic tool to identify territories at greatest risk for TB. Models should have broader scope to include social determinants so as to better inform policy to reduce inequality and achieving the goal of the End TB strategy.

## INTRODUCTION

Brazil is among the countries with the highest tuberculosis (TB) burden[1]. According to the latest report from the World Health Organization (WHO), Brazil identified more than 66,819 cases in 2020. This shows a decrease of 18% compared with data from the year 2019, but at the same time, there was an increase in TB-related mortality[1].

Although Brazil has committed to ending TB by 2050, the country has faced enormous challenges in achieving this ambitious goal. This is largely due to austerity measures that have cut social benefits to lower socio-economic groups as well as to a serious economic crisis brought on by the pandemic and government policies[2]. According to forecast models, the TB targets of the UN Sustainable Development Goals (SDGs) (which aims for a 90% reduction in TB deaths by 2030) are not likely to be achieved.

There is a broad agreement that progress in tuberculosis control will require not only investment in strengthening tuberculosis control programs, diagnostics, and treatment, but also action on the Social and structural Determinants of TB. The Structural factors include governing action, economic and social policies, and the position of power, prestige, and resources in accordance with the social level occupied by an individual, family, or community[3]. Intermediary determinants consist of material circumstances, psychosocial behavior, biological factors, and the health system[3]. Literature is replete with evidence of their influence in the context of TB,[4-9] but are without making distinctions on the dimensions of these determinants.

Mathematical modelling can be useful in exploring the contribution of health drivers to face the epidemic and browse through evidences to End TB strategies. However, current TB models have limitations, for example, a systematic review conducted by Pedrazzoli and colleagues showed that that few studies employing mathematical modeling have addressed underlying Social and Structural determinants, which makes this a knowledge gap[4]. Although many mathematical models have been used for forecasting TB, most of them tended to ignore the differences in variance and its asymmetry/imbalance and they retain almost exclusively the mean for explaining the outcome[10], which has revealed an important gap in knowledge.

Studies have provided results that are more satisfactory when alternative regressions techniques are applied. Generalized additive models for location, scale, and shape (GAMLSS) has allowed a more expanded approach where it worked with not only the mean (or location) but also all the parameters of the conditional distribution of outcome to be modeled as parametric/or nonparametric additive functions of independent variables and/or random-effects terms[11,12]. Therefore, here, we aimed to employ mathematical modeling to evaluate the impact of structural and intermediary social determinants on TB in Eastern Amazonia – Brazil.

## METHODS

### Study design

We used an ecological, mathematical modeling study design.

### The setting of the study

We conducted the study in Macapá, the capital of Amapá state, Eastern Amazonia (Fig 1). It has a population of about 398,204 inhabitants, and a demography density near 62.14 people per km2. It is the most populous city in Northern Amazonas State.

**Fig 1.**
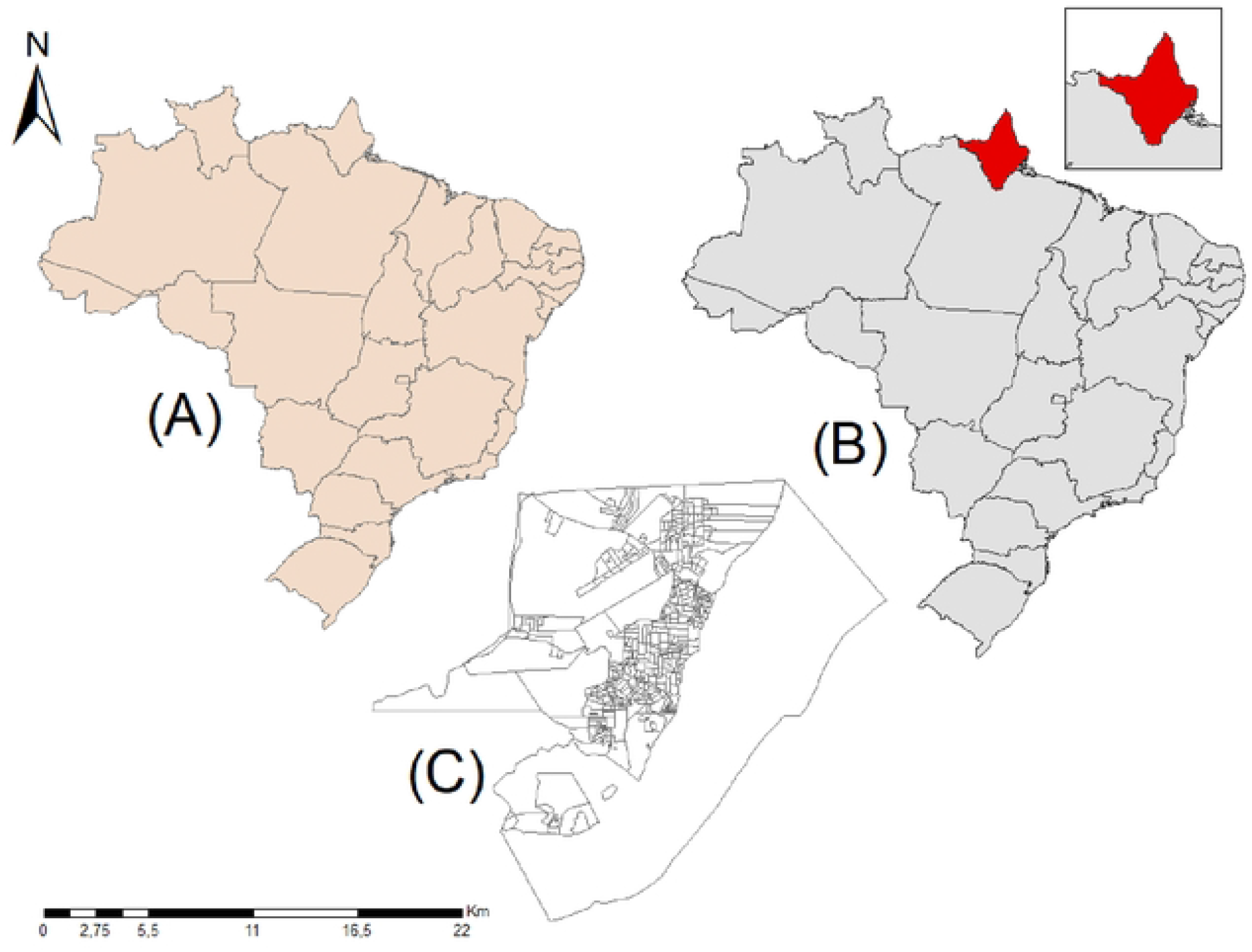
Map of the study setting.

Regarding TB, the city presented an incidence of 17.5 cases per 100,000 in 2020, which represented a decrease of 25% compared to the previous year’s data[13]. A problematic situation is that only 39.3% of the patients diagnosed with TB had confirmation from the bacteriological analysis (microscopy or GeneXpert MTB/RIF), and 47.3% did not have confirmation of the diagnosis of TB through a bacteriological test[14].

### The population of the study and criteria

The population was composed of all the cases diagnosed with tuberculosis and registered in the Notification of Notifiable Diseases System (SINAN) in the period from 2001 to 2017. We gathered socio-demographic data (age, gender, race/ color self-reported and years of study and occupation) and clinical information (type of case – new or retreatment; clinical form – pulmonary or extrapulmonary; coinfection with TB-HIV; alcoholism; mental disorders; comorbid TB-diabetes).

In accordance with the WHO guidelines, the diagnosis of TB refers to the recognition of an active case when a patient with *Mycobacterium tuberculosis complex* is identified from a clinical specimen, either by microscopy, culture, or a newer method such as a molecular line probe assay[15]. In Brazil, a pulmonary case with one or more initial sputum smear examinations positive for acid-fast bacilli (AFB) is also defined as a “case”[16]. Eventually, the diagnosis can be established only through clinical examination by a physician and X-ray; however, it is not recommended by the Brazilian sanitary authorities. New patients are defined as those who have no history of prior TB treatment or who have been treated for at least/ less one month. These cases should receive a regimen containing six months of rifampicin: 2HRZE/4HR[16].

### Unity of study analysis and variables

The unit of study analysis was 811 Urban Census Tract (UCT) contained in Macapá, collected from the Brazilian Institute of Geography and Statistics (IBGE). The UCT consists of the smallest territorial unit formed by a continuous area located in an urban area, with a defined size, number of households, and number of residents, used for Brazilian surveys and statistical research[17]. The information regarding UCT was obtained from the 2010 Brazilian Demographic Census, which collected data regarding both structural and intermediary determinants expressed by household conditions and characteristics of the territories (UCT). In Fig 2 is shown the variable under study.

**Fig 2.**
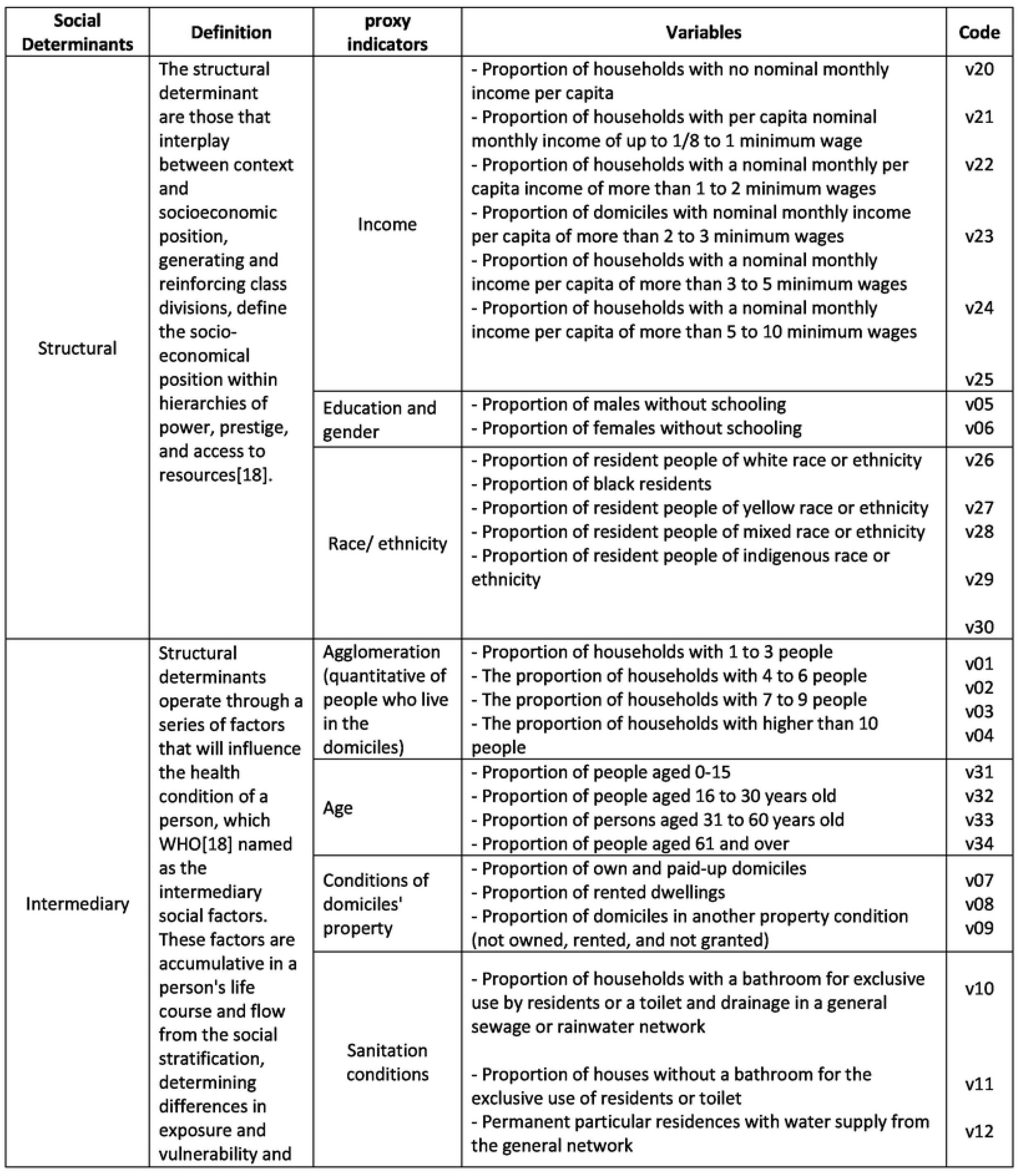
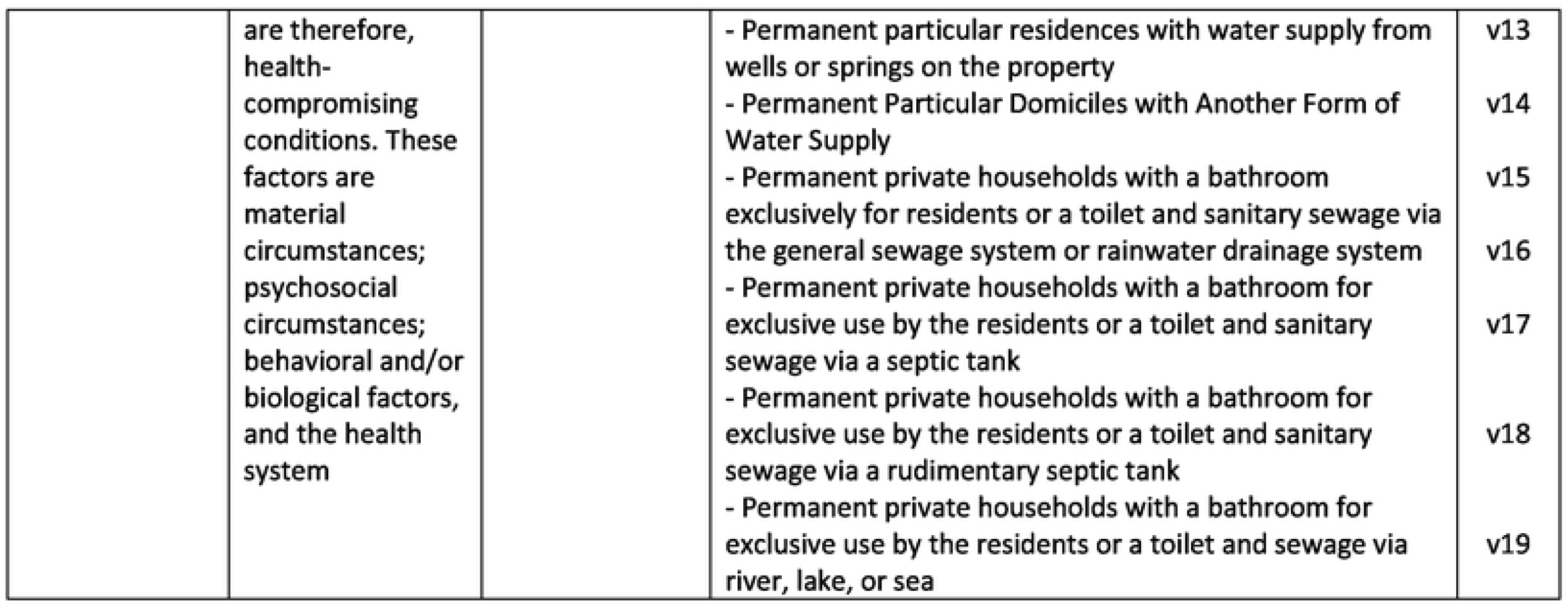
Structural and intermediary determinants selected for the study, Eastern Amazonia – Brazil.

### Analysis plan

Initially, the variables were analyzed by descriptive statistics. Additionally, for identifying the structural and intermediary determinants associated with TB, we used the model GAMLSS[11]. The reason for applying this model is because the modeling of the response variable (TB cases) did not follow a distribution of exponential family, and it showed heterogeneity in terms of distribution scale and shape. The response variable changed with the explanatory variables[19].

Since*y*^*T*^ *=* (*y*_1_, *…, y*_*n*_) is a vector of size n of the response variable with density function *f*(*y*_*i*_|*θ*^*i*^, where*θ*^*i*^ *=* (*θ*_1*i*_,*θ*_2*i*_ *θ*_3*i*_ *θ*_4*i*_) *=* (µ_*i*_,*σ*_*i*_ *υ*_*i*_ *τ*_*i*_), and let k = 1,2,3,4 e let*g*_*k*_(.) a monotone link function that relates the parameters to the independent variables from the equations:

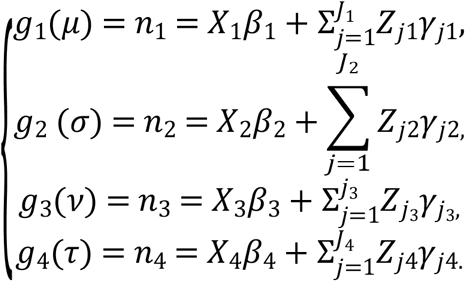

Where µ, σ, ν e τ are vectors of length n, 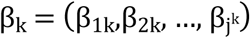 is a vector of lengthj^k^ eX_k_ is the delineation matrix of order n x j^k^. The function h_jk_

Non additive function of the independent variable X_k_ evaluated at x_jk_.

The selection of the dependent variable distribution was performed using the Generalized Akaike information criterion, defined by 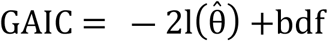, where 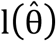the likelihood function, b is is a penalty parameter, and df denotes the degrees of freedom of the model[20]. For b = 2 we have the original Akaike information criterion (AIC). According to literature[21] all distributions falling in the GAMLSS class are presented.

The selection of the independent variables was made in 2 steps. In the first step, the presence of multicollinearity among the independent variables was evaluated. The multicollinearity assessment evaluates the entry of variables in the model that are highly correlated with each other. One of the most used measures is the Variance Inflation Factor, whose expression is defined by:

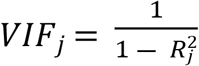

Where 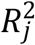 is the multiple correlation coefficient resulting from regressing*X*_*j*_ on the other p – 1 regressor. The higher the degree of dependence of*X*_*j*_ on the remaining regressors, the stronger the dependence and the higher the value of 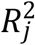. A VIF value > 5 was adopted as the cutoff point[12,22]. Since the independent variables are the same for both outcomes, the result of this analysis is also considered valid for both.

Additionally, we applied the stepwise method using the Generalized Akaike Information Criterion (GAIC) with k = 4[12] for the remaining variables from the first stage. After this analysis, The Double Poisson distribution (DPO) was selected in accordance with the AIC value (Table 3), where DPO (µ, σ) has the following probability density function[21].

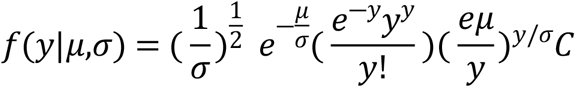

Where y = 0, 1, 2, …, ∞, µ > 0 and σ > 0, where C is a proportionality constant that is calculated numerically. The link function between the parameters and the independent variables is the log function, i.e., g_1 (µ)=log(µ) and g_2 (σ)=log(σ)[21].

We verified the model’s adequacy through the model’s diagnostic graphics: Fitted Values x Residuals, Order of Observations x Residuals, Distribution of Residuals, and the Quantile-Quantile plot (Q-Q plot). Additionally, the Shapiro-Wilk Normality test was applied to the model’s residuals to verify if they are fit with the Standard Normal distribution. We considered the dependent variable, the number of TB cases in each UCT, and the structural and intermediary determinants indicators as the independent variables (Table 1). These variable/ proxy indicators were selected in accordance with the theoretical framework defined in the study[4, 19].

**Table 1.** Socio-demographic and clinical-epidemiological profile of Tuberculosis cases, Eastern Amazonia (Brazil).

| Variables | N=1,730 (%) |
| --- | --- |
| Age (years) |  |
| 0 to 14 years | 74 (4.3) |
| 15 to 30 years old | 640 (37.0) |
| 31 to 59 years | 823 (47.6) |
| Above 60 years old | 175 (10.1) |
| Blank/Ignored | 18 (1.0) |
| Gender |  |
| Male | 704 (40,7) |
| Female | 1026 (59,3) |
| Years of study |  |
| Illiterate | 128 (7.4) |
| Elementary school complete | 112 (6.5) |
| Elementary School Incomplete | 803 (46.6) |
| High School Complete | 224 (12.9) |
| Higher Education Incomplete | 52 (3.0) |
| Higher education complete | 127 (7.3) |
| Blank/Ignored | 284 (16.4) |
| Registration types |  |
| New | 682 (39.4) |
| Relapse | 9 (0.5) |
| Retreatment after abandonment | 0 (0.0) |
| Retreatment | 1 (0.1) |
| Transfer | 1 (0.1) |
| After death | 26 (1.5) |
| Blank/Unknown | 1.011 (58.4) |
| Clinic form |  |
| Extrapulmonary | 236 (13.6) |
| Pulmonary | 1472 (85.1) |
| Pulmonary + Extrapulmonary | 18 (1.0) |
| Blank/Ignored | 4 (0.3) |
| Coinfection TB-HIV |  |
| No | 787 (45.5) |
| Yes | 77 (4.5) |
| Blank/Ignored | 866 (50.0) |
| Coinfection TB-Diabetes |  |
| No | 912 (52.8) |
| Yes | 94 (5.4) |
| Blank/Ignored | 724 (41.8) |
| Alcoholism |  |
| No | 896 (51.7) |
| Yes | 148 (8.6) |
| Blank/ Ignored | 686 (39.7) |
| Mental Disorders |  |
| No | 1001 (57.8) |
| Yes | 12 (0.7) |
| Blank/Ignored | 717 (41.5) |

We performed the GAIC selection of the independent variables considering only the linear effects. Additionally, we tested the inclusion of quadratic effects since in the scatter plots, the fitted curve (by the loess method = local polynomial regression) evidenced a possible quadratic relationship. To compare the two models, linear terms versus quadratic terms, we used the Likelihood Ratio Test (LR). Once we selected the fittest model, we estimated the Relative Increase, expressed in percentage, in the Average Number of Tuberculosis Cases through the expression*AR*(*β*) *=* [exp(*β*) ― 1] *∗* 100%. The R program version 4.1.1 through the GAMLSS library[23] was used to perform the data analysis for this part.

### Ethics approval and consent to participate

In compliance with Resolution 499/2012 of the National Health Council in Brazil, the study was authorized by the Municipal Health Secretariat of Macapá-Brazil and approved by the Research Ethics Committee from the University of São Paulo College of Nursing at Ribeirão Preto, which had the Certificate of Submission for Ethics Appreciation (CAAE) Number. 23043019.2.0000.5393. Consent to participate not applicable, because we work with secondary data. All participant identifiers were removed.

## RESULTS

We identified 1,730 patients diagnosed with TB. Table 1 shows the main characteristics of the patients, in which is observed an age ranging from 1 to 89 years (median = 44.5 years). Most were female (59.3%), aged 31 to 59 years (47.6%), blacks (67.9%), and with an education level of 5 to 8 incomplete years (18.7%). In terms of the clinical and epidemiological profile, cases were mostly characterized as new (39.4%) and pulmonary (85.1%) TB. Regarding comorbidities, we observed a TB-HIV prevalence of 4.5%; TB-Diabetes of 5.4%; Alcoholism at 8.6%; and mental illness at 0.7%. We observed an excess of cases numbers with ignored or blank data.

In Fig 3, evidences the spatial distribution of TB cases, considering the comorbid or health condition: TB-HIV coinfection mental disorder, diabetes or alcoholism.

**Fig 3.**
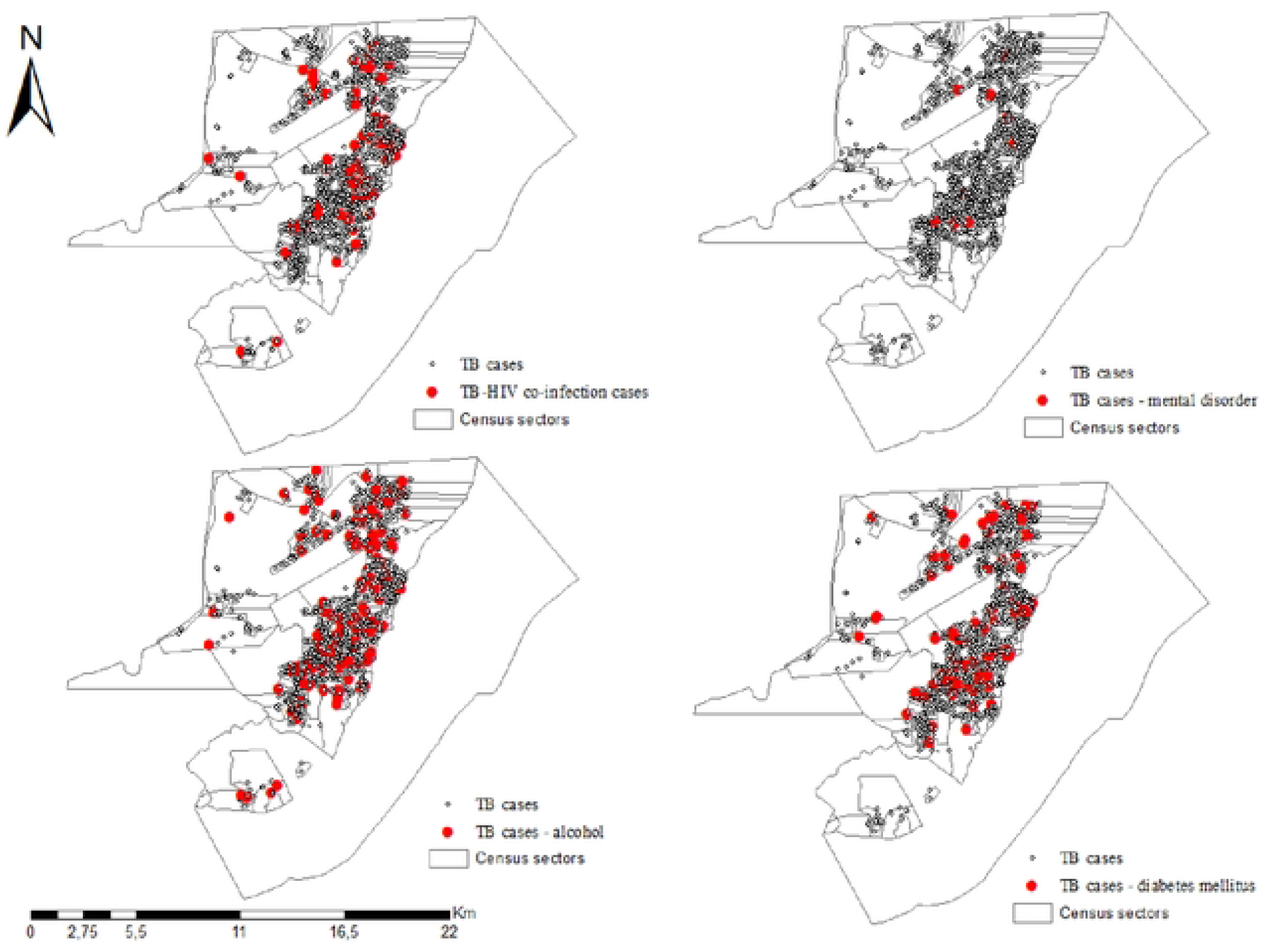
Spatial distribution of tuberculosis in the study, Eastern Amazonia.

In Table 2, we presented the main information obtained from the descriptive statistics. For the dependent variable, we considered the total of cases and the independents (proxy variables of structural and intermediary social determinants) the proportion of households in each UCT under those conditions characterized by the variable.

**Table 2.** Statistics descriptive of the variables defined in the study, Eastern Amazonia.

| **Variables** | **Mean** | **Standard Deviation** | **Minimum** | **Q0.25** | **Q0.5** | **Q0.75** | **Maximum** |
| --- | --- | --- | --- | --- | --- | --- | --- |
| Cases of TB per UCT * | 3.43 | 3.00 | 0.00 | 1.00 | 3.00 | 5.00 | 23.00 |
| v1 Proportion of households with 1 to 3 people | 42.32 | 7.56 | 19.15 | 37.77 | 42.21 | 47.19 | 76.81 |
| v2 Proportion of households with 4 to 6 people | 43.79 | 5.42 | 15.94 | 40.70 | 43.81 | 47.34 | 56.52 |
| v3 Proportion of households with 7 to 9 people | 10.82 | 4.07 | 0.00 | 8.28 | 10.17 | 12.86 | 29.79 |
| v4 Proportion of households with higher than 10 people | 3.08 | 2.19 | 0.00 | 1.60 | 2.70 | 4.04 | 14.89 |
| v5 Proportion of males without schooling | 18.33 | 7.28 | 3.28 | 13.09 | 17.60 | 22.73 | 44.06 |
| v6 Proportion of females without schooling | 17.14 | 6.82 | 2.33 | 12.33 | 16.12 | 21.38 | 43.90 |
| v7 Proportion of own and paid-up domiciles | 76.05 | 12.20 | 17.55 | 69.28 | 77.94 | 84.40 | 100.00 |
| v8 Proportion of rented dwellings | 16.70 | 9.72 | 0.00 | 9.45 | 15.32 | 22.65 | 67.11 |
| v9 Proportion of domiciles in another property' condition (not owned, rented, and not granted) | 0.19 | 0.67 | 0.00 | 0.00 | 0.00 | 0.00 | 10.20 |
| v10 Proportion of households with a bathroom for exclusive use by residents or a toilet and drainage in a general sewage or rainwater network | 65.63 | 14.40 | 9.09 | 56.64 | 65.60 | 75.00 | 100.00 |
| v11 Proportion of houses without a bathroom for the exclusive use of residents or toilet | 1.11 | 1.86 | 0.00 | 0.00 | 0.36 | 1.48 | 12.00 |
| v12 Permanent particular residences with water supply from the general network | 59.39 | 36.91 | 0.00 | 22.02 | 70.27 | 94.16 | 100.00 |
| v13 Permanent particular residences with water supply from wells or springs on the property | 36.73 | 34.79 | 0.00 | 4.49 | 25.65 | 70.83 | 100.00 |
| v14 Permanent Particular Domiciles with Another Form of Water Supply | 3.28 | 7.78 | 0.00 | 0.00 | 0.80 | 2.88 | 82.50 |
| v15 Permanent private households with a bathroom exclusively for residents or sanitary | 98.29 | 4.36 | 22.33 | 97.89 | 99.32 | 100.00 | 100.00 |
| v16 Permanent private households with exclusive use of bathroom or toilet and sanitary sewage via the general sewage network or rainwater drainage | 10.06 | 21.83 | 0.00 | 0.00 | 1.06 | 5.27 | 98.90 |
| v17 Permanent private households with a bathroom for exclusive use by the residents or a toilet and sanitary sewage via a septic tank | 18.73 | 26.59 | 0.00 | 0.88 | 6.02 | 25.09 | 100.00 |
| v18 Permanent private households with a bathroom for exclusive use by the residents or a toilet and sanitary sewage via a rudimentary septic tank | 49.23 | 33.94 | 0.00 | 14.18 | 54.88 | 81.91 | 99.36 |
| v19 Permanent private households with a bathroom for exclusive use by the residents or a toilet and sewage via river, lake, or sea | 17.28 | 28.16 | 0.00 | 0.00 | 1.09 | 21.06 | 100.00 |
| v20 Proportion of households with no nominal monthly income per capita | 4.07 | 4.83 | 0.00 | 1.31 | 2.68 | 5.06 | 38.64 |
| v21 Proportion of households with per capita nominal monthly income of up to 1/8 to 1 minimum wage | 58.02 | 20.22 | 7.63 | 43.61 | 60.00 | 73.71 | 96.09 |
| v22 Proportion of households with a nominal monthly per capita income of more than 1 to 2 minimum wages | 18.82 | 7.02 | 2.61 | 13.28 | 20.19 | 24.24 | 38.94 |
| v23 Proportion of domiciles with nominal monthly income per capita of more than 2 to 3 minimum wages | 7.75 | 5.63 | 0.00 | 3.14 | 6.85 | 11.41 | 36.36 |
| v24 Proportion of households with a nominal monthly income per capita of more than 3 to 5 minimum wages | 6.47 | 5.70 | 0.00 | 1.64 | 5.00 | 10.21 | 24.29 |
| v25 Proportion of households with a nominal monthly income per capita of more than 5 to 10 minimum wages | 3.93 | 4.57 | 0.00 | 0.71 | 2.24 | 5.55 | 23.25 |
| v26 Proportion of households with nominal monthly income per capita higher than 10 minimum wages | 1.20 | 2.05 | 0.00 | 0.00 | 0.42 | 1.42 |  |
| v27 Proportion of resident people of white race or ethnicity | 27.10 | 6.51 | 5.88 | 23.31 | 26.96 | 30.70 | 14.12 |
| v28 Proportion of black residents | 9.18 | 5.49 | 0.00 | 5.63 | 8.57 | 11.57 | 52.63 |
| v29 Proportion of resident people of yellow race or ethnicity | 1.19 | 1.34 | 0.00 | 0.26 | 0.80 | 1.80 | 39.60 |
| v30 Proportion of resident people of mixed race or ethnicity | 62.34 | 8.35 | 31.15 | 57.59 | 61.96 | 67.50 | 94.12 |
| v31 Proportion of resident people of indigenous race or ethnicity | 0.20 | 0.51 | 0.00 | 0.00 | 0.00 | 0.16 |  |
| v32 Proportion of people aged 0-15 | 32.74 | 7.21 | 9.44 | 27.46 | 32.64 | 38.28 | 6.77 |
| v33 Proportion of people aged 16 to 30 years old | 31.35 | 3.12 | 23.23 | 29.43 | 31.16 | 33.26 | 51.47 |
| v34 Proportion of persons aged 31 to 60 years old | 32.76 | 5.08 | 19.22 | 29.36 | 32.72 | 36.37 | 54.44 |
| v35 Proportion of people aged 61 and over | 5.16 | 2.93 | 0.87 | 3.21 | 4.16 | 6.10 | 47.73 |

In Table 3, we observed the results from the AIC criterion’s application for the selection of the most appropriate probability distribution considering the total number of TB cases, by which we can identify that the Double Poisson distribution (DPO) presented the best result (the highest AIC).

**Table 3.**
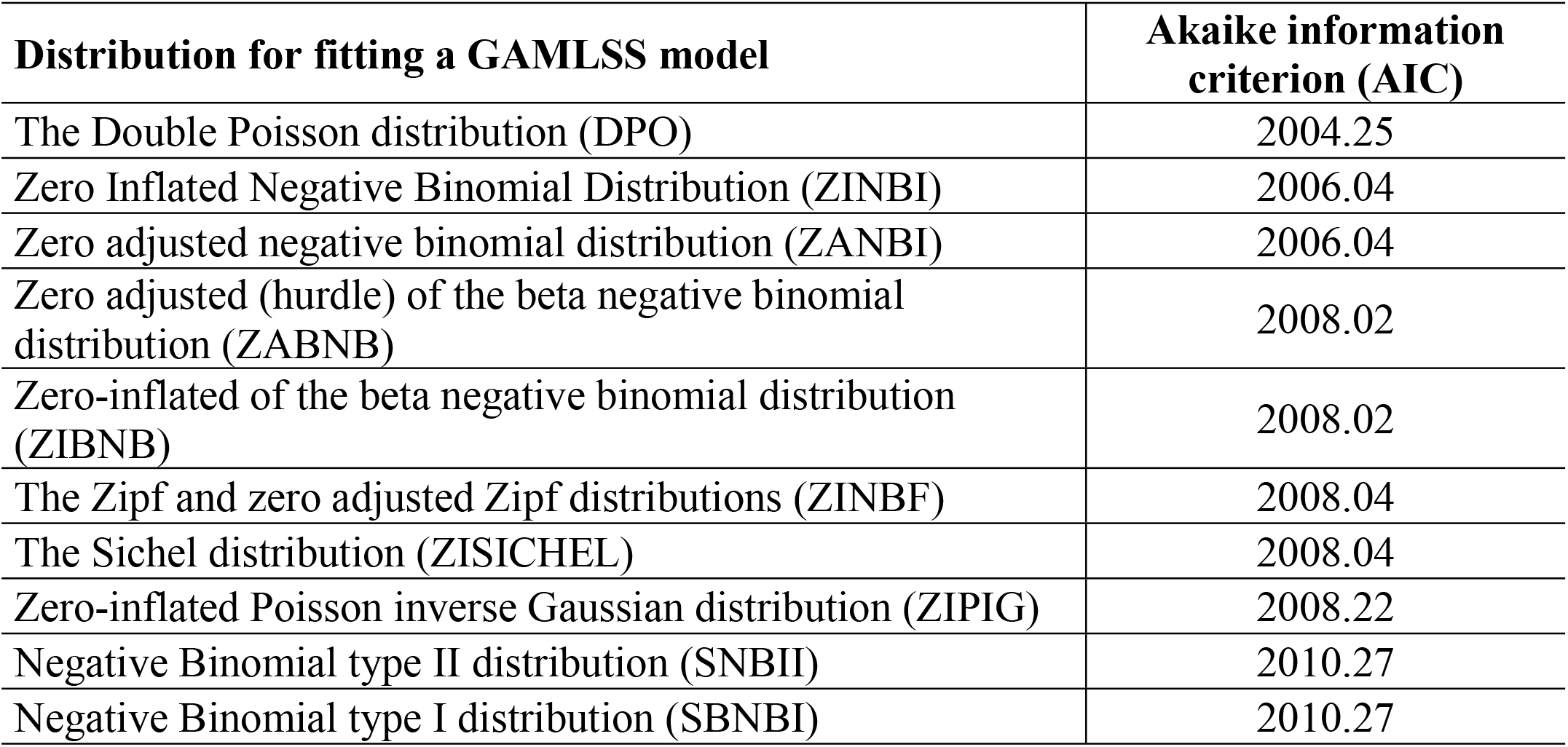
The main distributions for fitting a GAMLSS model selected in accordance with the Akaike Information Criterion value, Eastern Amazonia (Brazil).

In Fig 4, we have the superposition of the DPO density over the data distribution. The application of the Shapiro-Wilk normality test on the model’s residuals shows the adequacy of the fit (W = 0.9967; p-value = 0.5245).

**Fig 4.**
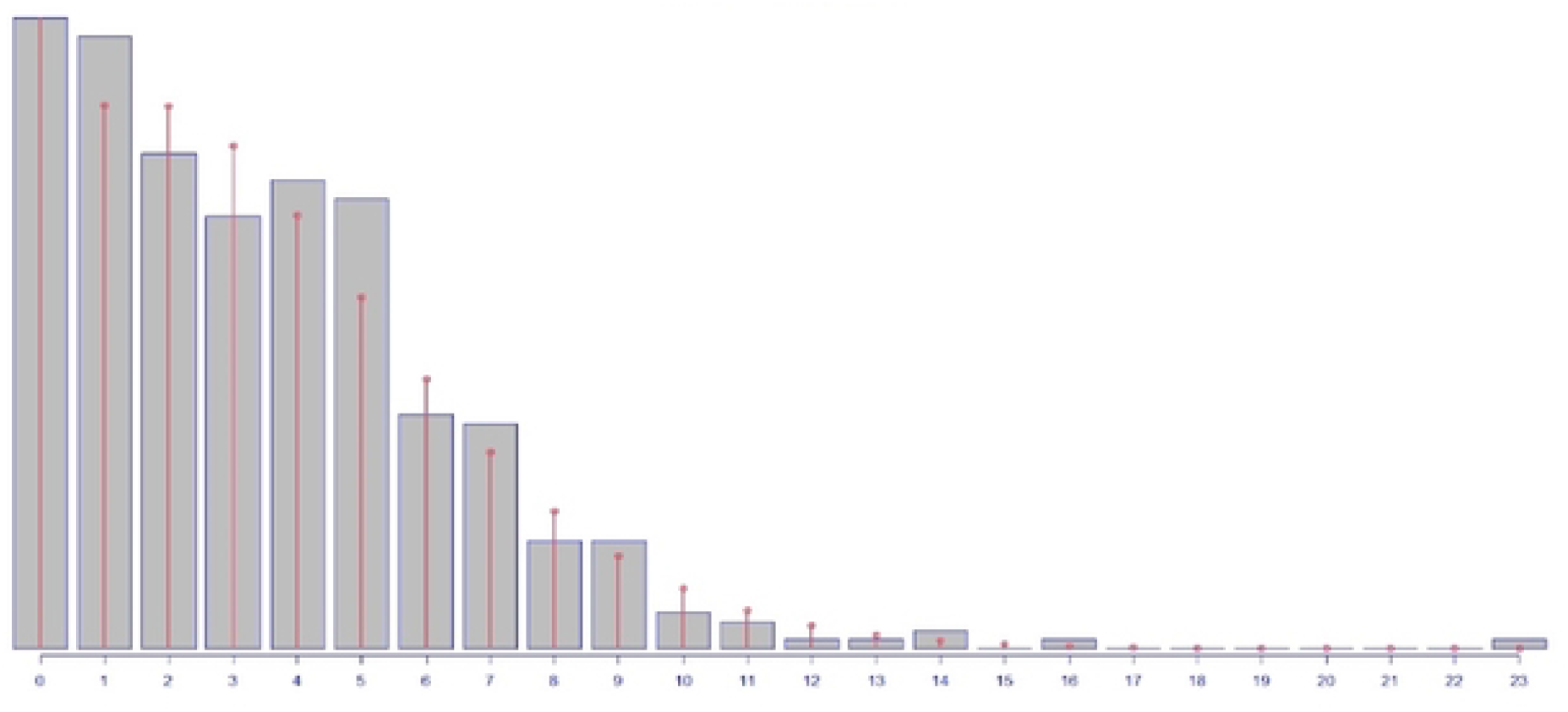
Histograms of Tuberculosis cases with fit to the transformed Double Poisson distribution, Eastern Amazonia (Brazil).

In Table 4, we included the statistics obtained from the modeling analysis, where we identify the complete model (saturated) selected through GAIC (no outlier) with quadratic terms and no quadratic outlier (v33 and v19). These variables represent factors associated with territories specifically deficient or absent of a sewage disposal system and the prevalence of younger people (16 to 30 years old) or older (higher than 61 years old). After the application of LR, the model with the quadratic terms was more fit when compared to the linear model. In the table, v19^1 is the linear term of v19 and v19^2 is the quadratic term of v19. The same explanation should be considered in v33.

**Table 4.**
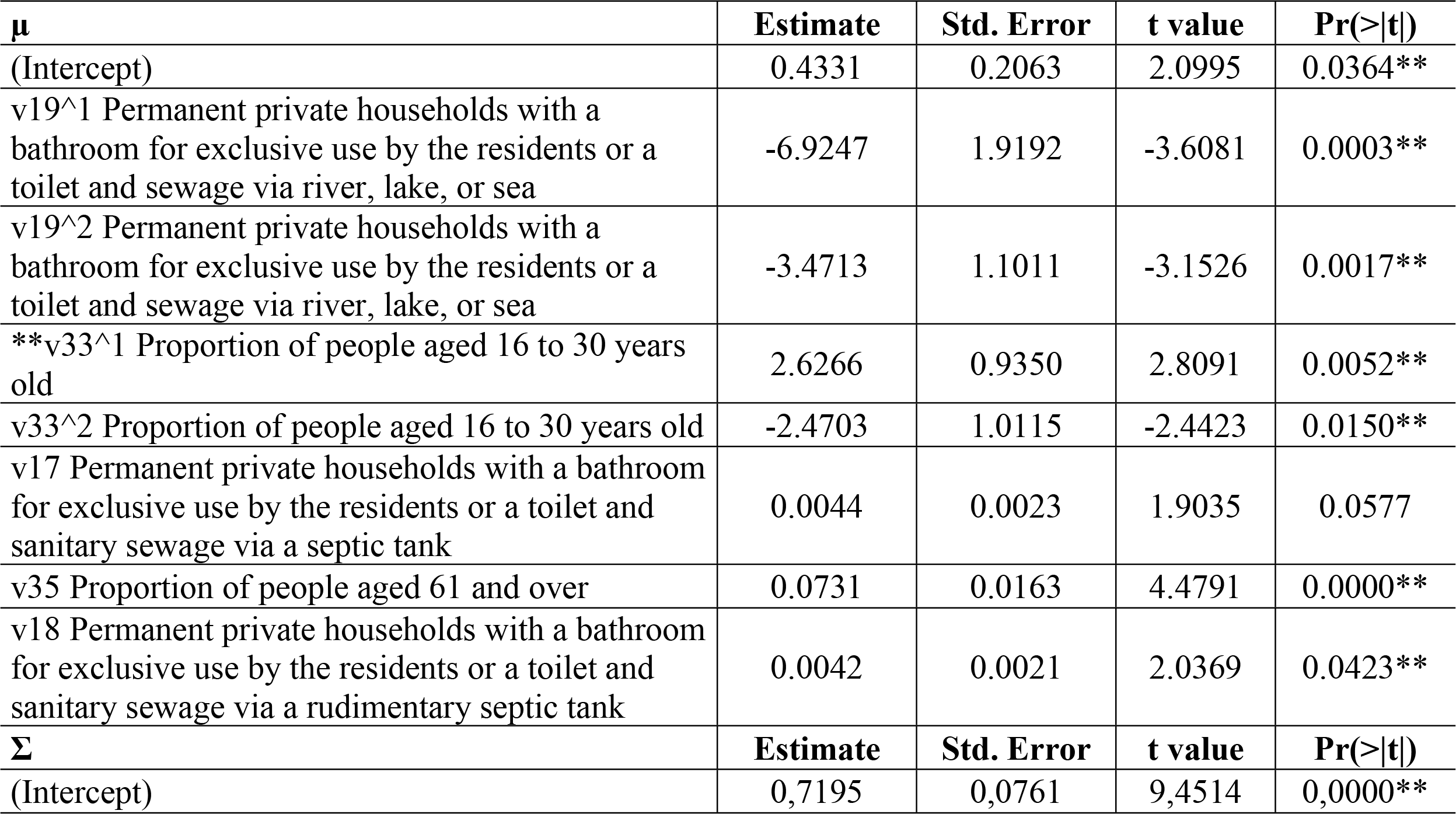
Model of the structural and intermediary social determinants associated with community TB infection in Eastern Amazonia (Brazil). *Shapiro-Wilk normality test; data: residuals (ajuste_o2c) W = 0.99681, p-value = 0.5654; Likelihood Ratio Test for nested GAMLSS models; (No check whether the models are nested is performed). Null model: deviance= 1884.811 with 7 deg. of freedom Alternative model: deviance= 1866.812 with 9 deg. of freedom; LRT = 17.99909 with 2 deg. of freedom and p-value= 0.0001234661 **p<0.05

| $\mu$ | Estimate | Std. Error | t value | Pr(>\|t\|) |
| --- | --- | --- | --- | --- |
| (Intercept) | 0.4331 | 0.2063 | 2.0995 | 0.0364** |
| v19^1 Permanent private households with a bathroom for exclusive use by the residents or a toilet and sewage via river, lake, or sea | -6.9247 | 1.9192 | -3.6081 | 0.0003** |
| v19^2 Permanent private households with a bathroom for exclusive use by the residents or a toilet and sewage via river, lake, or sea | -3.4713 | 1.1011 | -3.1526 | 0.0017** |
| **v33^1 Proportion of people aged 16 to 30 years old | 2.6266 | 0.9350 | 2.8091 | 0.0052** |
| v33^2 Proportion of people aged 16 to 30 years old | -2.4703 | 1.0115 | -2.4423 | 0.0150** |
| v17 Permanent private households with a bathroom for exclusive use by the residents or a toilet and sanitary sewage via a septic tank | 0.0044 | 0.0023 | 1.9035 | 0.0577 |
| v35 Proportion of people aged 61 and over | 0.0731 | 0.0163 | 4.4791 | 0.0000** |
| v18 Permanent private households with a bathroom for exclusive use by the residents or a toilet and sanitary sewage via a rudimentary septic tank | 0.0042 | 0.0021 | 2.0369 | 0.0423** |
| $\Sigma$ | Estimate | Std. Error | t value | Pr(>\|t\|) |
| (Intercept) | 0,7195 | 0,0761 | 9,4514 | 0,0000** |

Fig 5 presents the graph of predicted values containing the values of the variables with quadratic terms v19 and v33. Considering that variables v17, v18 and v35 are equal in terms of their respective medians, in other words, v17 = 18.74%, v18 = 49.35% and v35 = 5.17%). In this case, if we consider that v19=10%, v33=20%, the expected average number of TB cases is 0.7367; When we consider v19 = 20% and v33 = 30%, the expected average number of cases is 3.7824; and if we consider v19 = 30% and v33 = 40%, the average expected number of cases is 2.86. Fig 6 has been added as supplementary material for predicting the number of cases in accordance with the ranged value of v19 and v33.

**Fig 5.**
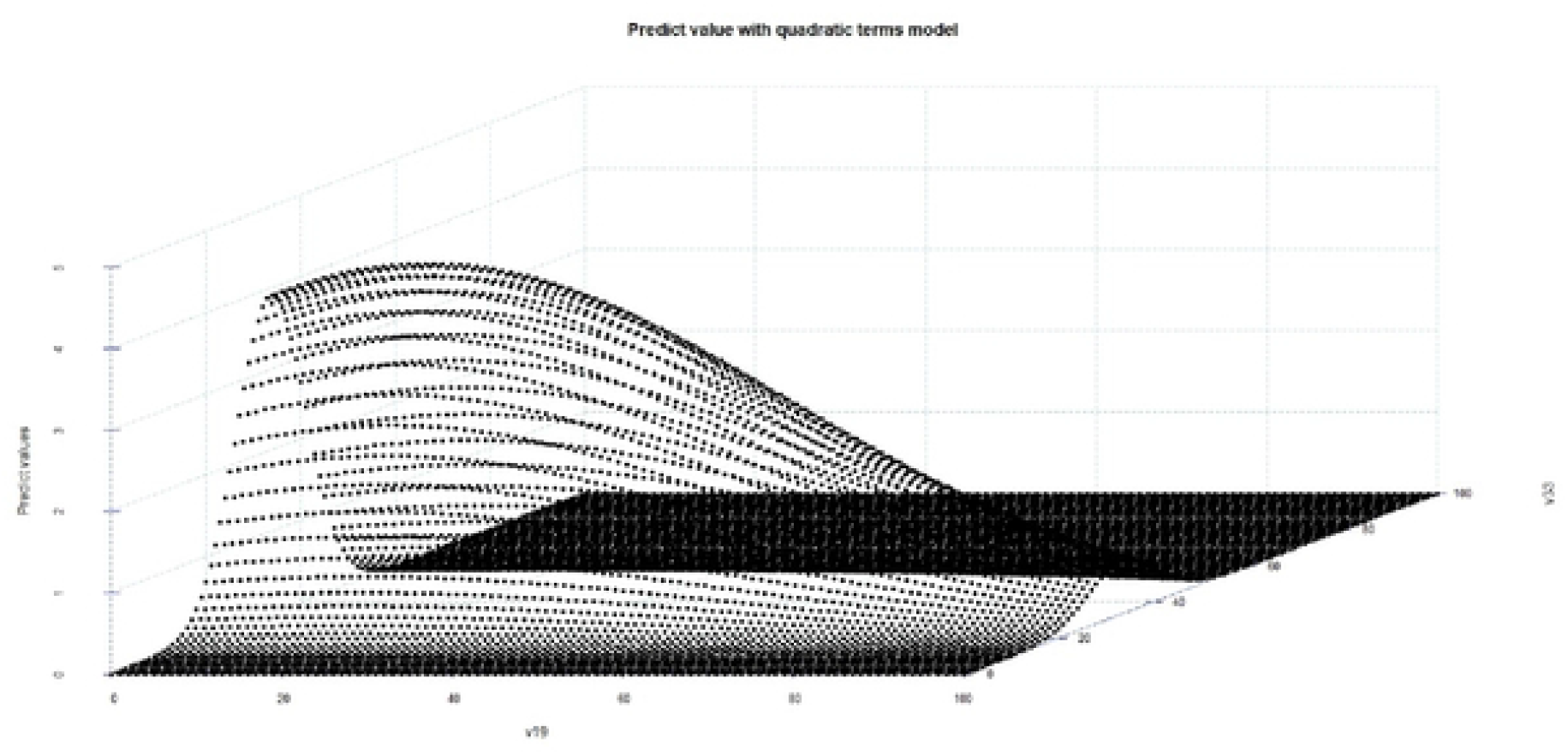
Graph with predicted values obtained from the modeling with quadratic terms, Eastern Amazonia (Brazil).

**Fig 6.**
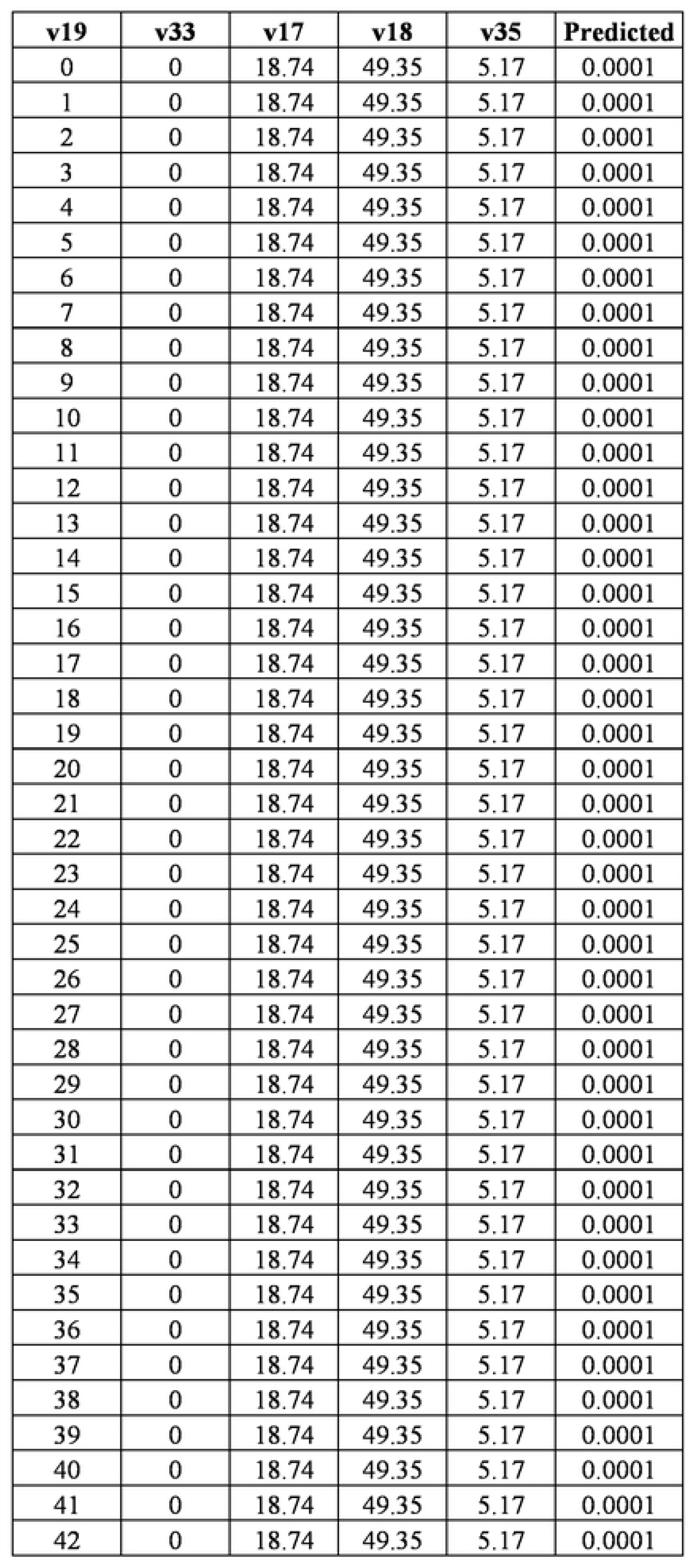
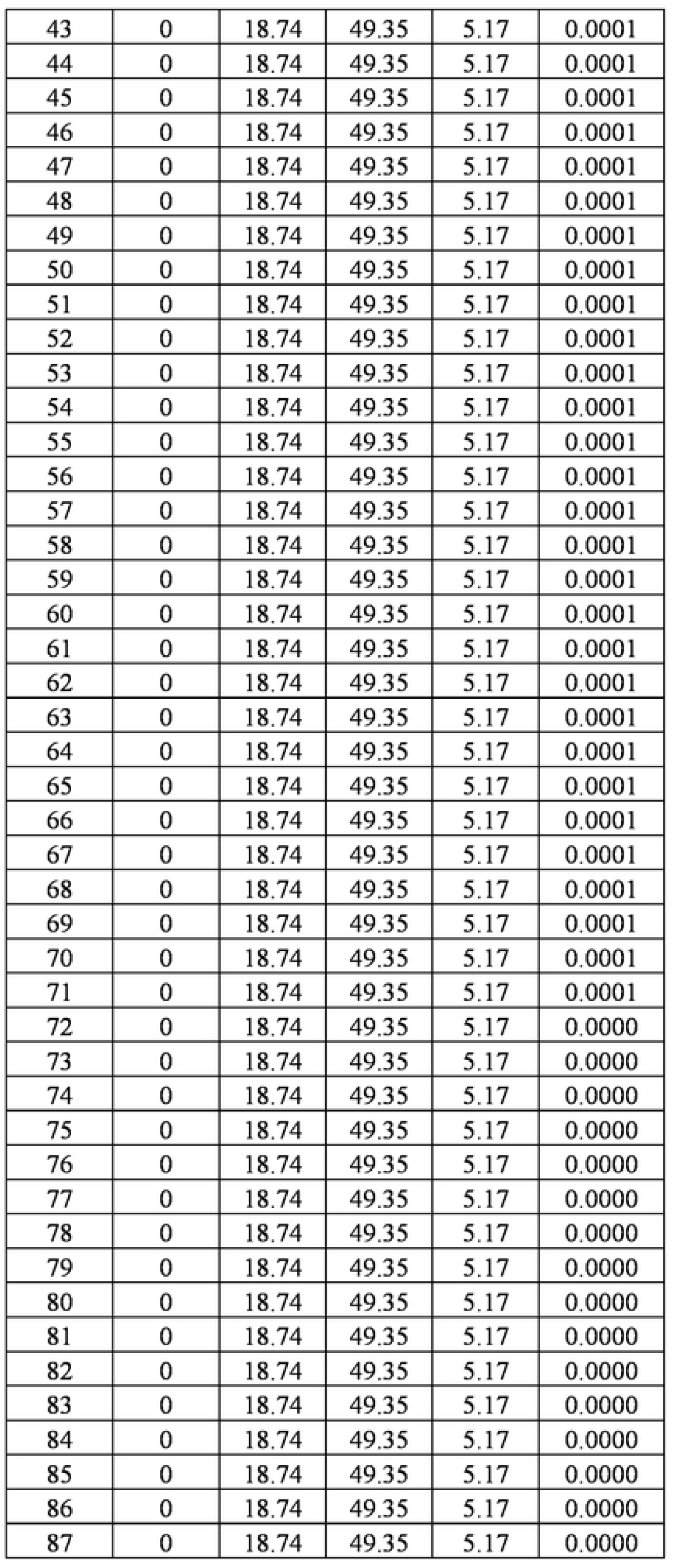
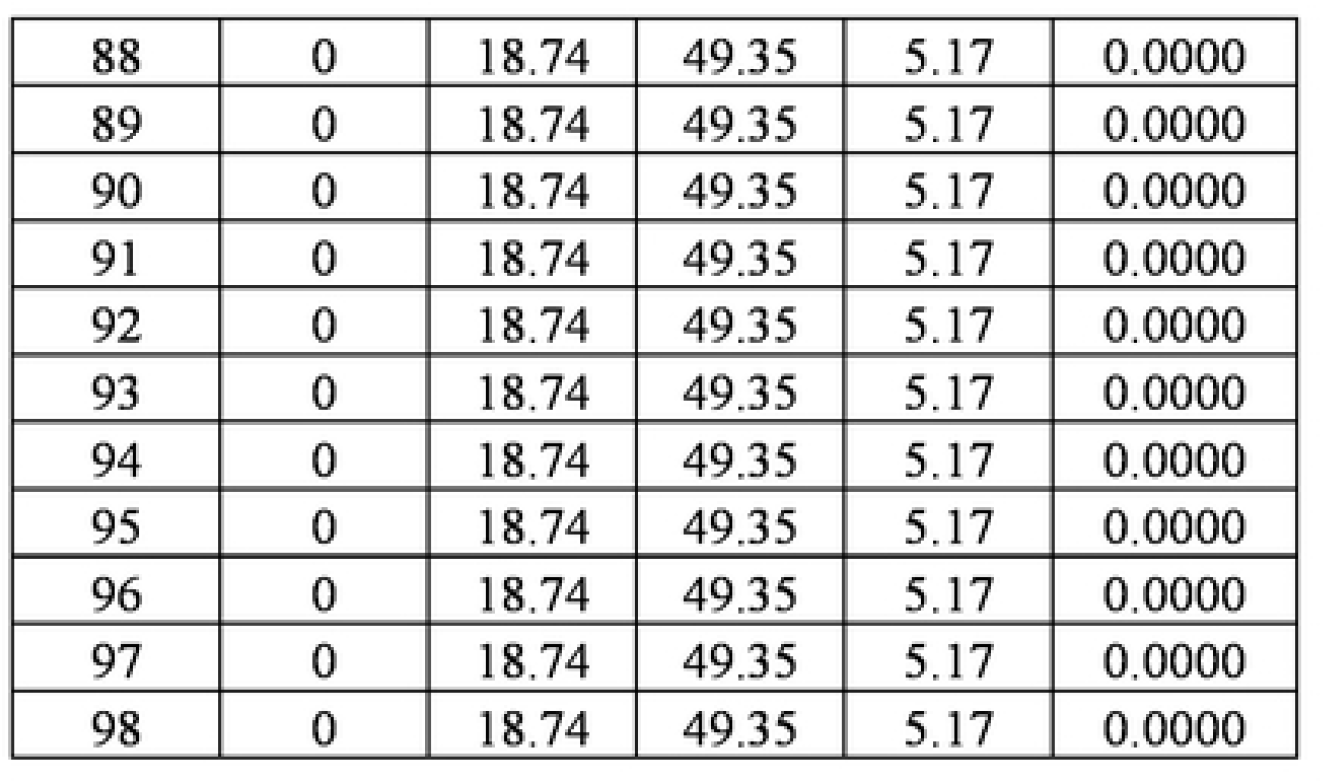
Predicting the number of cases according to the range value of v19 and v33.

## DISCUSSION

We found evidence that GAMLSS is superior when compared to the other techniques applied for studying the social determinants[12]. The first potentiality is because the GAMLSS estimates the complete conditional distribution, which means that the response variable can be better estimated by a probability distribution such as the gamma or lognormal distribution[24]. Another potentiality or advantage is that all resources are available through R packages, where it is allowed to adjust more than 50 different kinds of distributions.

Through our findings, we identified the structural determinants (represented by age) and intermediary (expressed by sanitary conditions of the environment such as treated sewage). The literature has evidenced that people of economically active age who are males were more commonly affected by TB, which might be related to our findings[25].

According to the national data, nearly 8.5% of TB cases that were diagnosed in 2017 were in people 19 or younger[25]. In Brazil, TB in adolescents has long remained a hidden pandemic and still continues to be neglected. TB in adolescents needs to be considered a sentinel event since it is related to a recent infection through contact with a bacilliferous adult[25].

Another issue that may be associated with our findings is a situation revealed by a recent IBGE survey, which concluded that the country has 14.8 million unemployed people, which represents 14.7% of the economically active population. However, this rate is even higher among young people; in the 14 to 17 age group, 46% are looking for work; and from 18 to 24 years old, unemployment affects 31% of people, which takes these people into an extreme social vulnerability zone. The study also evidenced that long-term unemployment is characterized by the predominance of people between the ages of 17 and 29. This data is from Brazil, but when it is considered data from Eastern Amazonia, this situation can become even more serious[26].

We also found the association with territories with a predominance of elderly people (age older than 61 years old), and the literature has shown that the elderly are more vulnerable to falling into poverty when compared to the other groups[27]. This is because the elderly have a lesser chance of recovering from a negative income shock, and they have difficulties (re)entering the labor market because productivity and employability decline with age from around the age of 60. As a result of the difficulty of recovering from negative income shocks among the elderly, poverty tends to be a more permanent feature than with other groups in the society[27], which makes them vulnerable to poverty as well as to the TB determinants[28].

A study from Eastern Amazonia has shown a growth of TB among the elderly, which is related to the deterioration of social conditions because of austerity politics adopted in Brazil, mainly related to the weakness in social security in ensuring social protection to the elderlies, and their difficulty in getting good food, quality of life and improved house conditions[29].

We also observed that the context favorable for sickness due to TB is associated with access to basic sanitation. This is another serious situation observed in developing countries; according to literature, more than 2 billion people worldwide do not have access to basic sanitation, which represents more than 25% of the world’s population[30]. According to a study that assessed the access to basic sanitation in Brazil, almost 90% of the residents of Macapá do not have access to the sewage network, and as in the whole country, Macapá invests less than 30% of their financial resources picked up with taxes[31].

Investments in terms of better housing conditions and basic sanity is a condition essential for the quality of life of people as well as for avoiding neglected diseases such as TB. The Sustainable Development Goals, also known as the Global Goals, seek to reconcile economic growth, environmental balance, and social progress, ensuring that all people have the same opportunities and can lead a better life without compromising the planet[32]. It is among its goals by 2030 to achieve universal and equitable access to basic sanitation and clean water; however, achieving these goals is a challenge in Brazil, mainly in Eastern Amazonia.

Eastern Amazonia has the largest percentage of its territory set aside for integral natural protection through the so-called Legally Allocated Areas, and contradictorily, has almost half of its population living below the poverty line, with 45.9% of people having one full meal every three days. A large part living in subhuman conditions in hangover areas, worsening the pictures of violence, suicide, and public health, while leaving 20.2% of its entire labor force unemployed. Therefore, all these aspects contributed a lot to the sickness of the community due to TB. It is important to find strategies not only for prevention, diagnosis, treatment, and recovery / rehabilitation, but to fundamentally find solutions sustainable and balanced with the environment in Amazonia.

Therefore, politics addressed to ensure sustainability is quite important for bringing better conditions of life to the people who live in the region. Historically, the poverty in the region became evident from the decolonization process with the formation of a dependent labor market in extremely precarious conditions. New strategies are important and effective public policies aimed at social justice to the specificities of the Amazon region and not just an attempt to solve social inequalities with compensatory policies such as the *Bolsa Familia*[33].

The study advances knowledge by evidencing the structural and intermediary social determinants of TB in Eastern Amazonia. However, the model only included cases of TB. It would be interesting to advance with other aspects of the disease through mathematical models such as infection and confirm if they are the same as the disease[4]. We used the GAMLSS that evidenced important aspects for understanding the context of TB and how the structural and intermediary determinants impacted on communities burdened by TB, where it was possible to estimate the number of cases for each territory or UCT under study.

We used a more usual approach, which did not assume the existence of the constraint, which may be a potential limitation of the study. This is the first study applied in Eastern Amazonia that confirms its novelty and originality. Unfortunately, Brazil has not carried out its Demographic Census yet, mainly because of the budget shortfall. Therefore, the social situation identified may have worsened, mainly due to the COVID-19 pandemic, which means that the goal End TB by 2050 seems more distant than we think.

## Data Availability

All relevant data are within the manuscript and its Supporting Information files (S1_Dataset).

## Acknowledgements

The authors greatly appreciate the outstanding and committed study assistance of Health Bureau of Mapacá – AP Brazil. We thank Jason Farley and Nancy Reynolds for critically reading the manuscript. Moreover, we thank CAPES and CNPq for contributing to the study.

## Author Contributions

**Conceptualization:** Clóvis Luciano Giacomet, Antônio Carlos Vieira Ramos, Heriederson Sávio Dias Moura, Thaís Zamboni Berra, Ricardo Alexandre Arcêncio.

**Data curation:** Clóvis Luciano Giacomet, Antônio Carlos Vieira Ramos, Thaís Zamboni Berra, Jonas Bodini Alonso.

**Formal analysis:** Clóvis Luciano Giacomet, Antônio Carlos Vieira Ramos, Thaís Zamboni Berra, Jonas Bodini Alonso.

**Funding acquisition:** Clóvis Luciano Giacomet, Heriederson Sávio Dias Moura, Ricardo Alexandre Arcêncio.

**Methodology:** Clóvis Luciano Giacomet, Antônio Carlos Vieira Ramos, Thaís Zamboni Berra, Felipe Mendes Delpino, Jonas Bodini Alonso, Ricardo Alexandre Arcêncio.

**Software:** Jonas Bodini Alonso, Ricardo Alexandre Arcêncio.

**Supervision:** Clóvis Luciano Giacomet, Ricardo Alexandre Arcêncio.

**Validation:** Clóvis Luciano Giacomet, Antônio Carlos Vieira Ramos, Thaís Zamboni Berra, Jason E. Farley, Nancy R. Reynolds, Jonas Bodini Alonso, Ricardo Alexandre Arcêncio.

**Writing – original draft:** Clóvis Luciano Giacomet, Antônio Carlos Vieira Ramos, Heriederson Sávio Dias Moura, Thaís Zamboni Berra, Yan Mathias Alves, Felipe Mendes Delpino, Jonas Bodini Alonso, Ricardo Alexandre Arcêncio

**Writing – review & editing:** Clóvis Luciano Giacomet, Antônio Carlos Vieira Ramos, Heriederson Sávio Dias Moura, Thaís Zamboni Berra, Yan Mathias Alves, Felipe Mendes Delpino, Jason E. Farley, Nancy R. Reynolds, Jonas Bodini Alonso, Titilade Kehinde Ayandeyi Teibo, Ricardo Alexandre Arcêncio.

## Supporting information

S1 – Dataset

## REFERENCES

1. World Health Organization. Global tuberculosis report 2021, 2021. Accessed: Dec 10, 2021. Available from: https://apps.who.int/iris/handle/10665/346387.

2. United Nations. World Social Report 2020: Inequality in a Rapidly Changing World, 2020. Accessed Dec 10, 2021. Available from: https://www.un-ilibrary.org/economic-and-social-development/world-social-report-2020_7f5d0efc-en.

3. Silva S, Arinaminpathy N, Atun R, Goosby E, Reid M. Economic impact of tuberculosis mortality in 120 countries and the cost of not achieving the Sustainable Development Goals tuberculosis targets: a full-income analysis. Lancet Glob Health. 202;9(10): e1372–9. doi: 10.1016/S2214-109X(21)00299-0.

4. Pedrazzoli D, Boccia D, Dodd PJ, Lönnroth K, Dowdy DW, Siroka A, et al. Modelling the social and structural determinants of tuberculosis: opportunities and challenges. Int J Tuberc Lung Dis Off J Int Union Tuberc Lung Dis. 2017;21(9): 957–64. doi: 10.5588/ijtld.16.0906.

5. Moreira ASR, Kritski AL, Carvalho ACC. Social determinants of health and catastrophic costs associated with the diagnosis and treatment of tuberculosis. J Bras Pneumol. 2020;46(05): e20200015. doi: 10.36416/1806-3756/e20200015.

6. Duarte R, Lönnroth K, Carvalho C, Lima F, Carvalho ACC, Muñoz-Torrico M, et al. Tuberculosis, social determinants and co-morbidities (including HIV). Pulmonology 2018;24(2): 115–119. doi: 10.1016/j.rppnen.2017.11.003.

7. Zille AI, Werneck GL, Luiz RR, Conde MB. Social determinants of pulmonary tuberculosis in Brazil: an ecological study. BMC Pulm Med. 19(1): 87. doi: 10.1186/s12890-019-0855-1.

8. Kilabuk E, Momoli F, Mallick R, Van Dyk D, Pease C, Zwerling A, et al. Social determinants of health among residential areas with a high tuberculosis incidence in a remote Inuit community. J Epidemiol Community Health. 73(5): 401–406. doi: 10.1136/jech-2018-211261.

9. Wingfield T, Tovar MA, Datta S, Saunders MJ, Evans CA. Addressing social determinants to end tuberculosis. Lancet. 391(10126): 1129–1132. doi: 10.1016/S0140-6736(18)30484-7.

10. Rigby RA, Stasinopoulos DM. Generalized additive models for location, scale and shape. J R Stat Soc Ser C Appl Stat. 2005;54(3): 507–54. doi: 10.1111/j.1467-9876.2005.00510.x.

11. De Bastiani F, Rigby RA, Stasinopoulous DM, Cysneiros AHMA, Uribe-Opazo MA. Gaussian Markov random field spatial models in GAMLSS. J Appl Stat. 45(2018): 168–86. doi: 10.1080/02664763.2016.1269728.

12. Giacomet CL, Santos MS, Berra TZ, Alves YM, Alves LS, Costa FBP, et al. Temporal trend of tuberculosis incidence and its spatial distribution in Macapá – Amapá. Rev Saude Publica. 55: 96. doi: 10.11606/s1518-8787.2021055003431.

13. Santana JLA, Magrini AT, Ribeiro LA, Rigelli RH, Araujo MHM. Adesão ao tratamento da tuberculose no Amapá: Um quinquênio de análise epidemiológica. Rev Científica Multidiscip Núcleo Conhecimento. 10: 69–87. doi: 10.32749/nucleodoconhecimento.com.br/saude/tratamento-da-tuberculose.

14. World Health Organization. Meeting report of the WHO expert consultation on drug-resistant tuberculosis treatment outcome definitions, 17–19 November 2020. Accessed: Dec 13, 2021. Available from: https://apps.who.int/iris/handle/10665/340284.

15. Brasil. Ministério da Saúde. Secretaria de Vigilância em Saúde. Departamento de Vigilância das Doenças Transmissíveis. Manual de Recomendações para o Controle da Tuberculose no Brasil, 2019; 2a ed. atualizada, 364. Accessed: Dec 13, 2021. Available from: https://bvsms.saude.gov.br/bvs/publicacoes/manual_recomendacoes_controle_tuberculose_brasil_2_ed.pdf.

16. Boing AF, Boing AC, Subramanian SV. Inequalities in the access to healthy urban structure and housing: an analysis of the Brazilian census data. Cad Saúde Pública. 37(6): e00233119. doi: 10.1590/0102-311X00233119.

17. Solar O, Irwin A. A conceptual framework for action on the social determinants of health, Social Determinants of Health Discussion Paper 2 (Policy and Practice). Geneva, Switzerland: World Health Organization. 2010. Accessed: Dec 10, 2021. Available from: https://apps.who.int/iris/handle/10665/44489.

18. Florencio LA. Engenharia de avaliações com base nos modelos GAMLSS. Master’s Thesis, Federal University of Pernambuco, 2010. Available from: https://repositorio.ufpe.br/handle/123456789/6227.

19. Akaike H. A new look at the statistical model identification. IEEE Trans Autom Control. 19(6): 716–23. doi: 10.1109/TAC.1974.1100705.

20. Stasinopoulos M, Rigby B, Akantziliotou C. Instructions on how to use the gamlss package in R, 2008. Accessed: Dec 14, 2021. Available from: http://www.gamlss.com/wp-content/uploads/2013/01/gamlss-manual.pdf.

21. Myers RH, Montgomery DC, Anderson-Cook CM. Response surface methodology: process and product optimization using designed experiments. Vanderbilt University, Hoboken, New Jersey: Wiley, 2016. Accessed: Dec 21, 2021. Available from: https://catalog.library.vanderbilt.edu/discovery/fulldisplay/alma991043342547203276/01VAN_INST:vanui.

22. Stasinopoulos DM, Rigby RA. Generalized Additive Models for Location Scale and Shape (GAMLSS) in R. Journal of Statistical Software. 23(7): 1–46. https://doi.org/10.18637/jss.v023.i07.

23. Jesus FB, Lima FCA, Martins CBG, Matos KF, Souza SPS. Vulnerabilidade na adolescência: a experiência e expressão do adolescente. Rev Gaúcha Enferm. 32(2): 359–67. doi: 10.1590/S1983-14472011000200021.

24. Shelby T, Meyer AJ, Ochom E, Turimumahoro P, Babirye D, Katamba A, et al. Social determinants of tuberculosis evaluation among household contacts: a secondary analysis. Public Health Action. 8(3): 118–123. doi: 10.5588/fa.18.0025.

25. Santos BA, Cruz RPS, Lima SVMA, Santos AD, Duque M, Araújo KCGM, et al. Tuberculose em crianças e adolescentes: uma análise epidemiológica e espacial no estado de Sergipe, Brasil, 2001-2017. Ciênc Saúde Coletiva. 25(8): 2939–48. doi: 10.1590/1413-81232020258.25692018.

26. Brasil. Instituto Brasileiro de Geografia e Estatísticas. Pesquisa aponta que os jovens são os mais afetados pelo desemprego, 2021. Accessed: Jan 21, 2021. Available from: https://agenciabrasil.ebc.com.br/radioagencia-nacional/economia/audio/2021-08/pesquisa-aponta-que-os-jovens-sao-os-mais-afetados-pelo-desempregohtml.

27. Travassos GF, Coelho AB, Arends-Kuenning MP. The elderly in Brazil: demographic transition, profile, and socioeconomic condition. Rev Bras Estud Popul. 37: 1–27. doi: 10.20947/S0102-3098a0129.

28. Negin J, Abimbola S, Marais BJ. Tuberculosis among older adults – time to take notice. Int J Infect Dis. 32: 135–7. doi: 10.1016/j.ijid.2014.11.018.

29. Mesquita CR, Lima KVB, Souza e Guimarães RJP, Santos BO, Rodrigues LHA, Costa RJF, et al. Análise retrospectiva de casos de tuberculose em idosos. Rev Bras Em Promoção Saúde 2021;34: 1117. doi: 10.5020/18061230.2021.11117.

30. Swe KT, Rahman MM, Rahman MS, Teng Y, Abe SK, Hashizume M, et al. Impact of poverty reduction on access to water and sanitation in low- and lower-middle-income countries: country-specific Bayesian projections to 2030. Trop Med Int Health. 26(7): 760–774. doi: 10.1111/tmi.13580.

31. Instituto Trata Brasil. Ranking do Saneamento 2021, 2021. Accessed: Jan 19, 2022. Available from: https://tratabrasil.org.br/images/estudos/Ranking_saneamento_2021/Ranking_do_Saneamento_2021_-_tabela_das_100_maiores_cidades_do_Brasil_.pdf.

32. Duran DC, Artene A, Gogan LM, Duran V. The Objectives of Sustainable Development -Ways to Achieve Welfare. Procedia Econ Finance. 26(2015): 812–7. doi: 10.1016/S2212-5671(15)00852-7.

33. Rolim DC. A pobreza e a riqueza na região amazônica e a contribuição da política de assistência social: o Estado do Amazonas em foco. VII Jornada Internacional Políticas Públicas. Universidade Federal do Maranhão. Centro de Ciências Sociais. Programa de Pós-Graduação em Políticas Públicas 2015:13. Accessed: Jan 19, 2022. Available from: http://www.joinpp.ufma.br/jornadas/joinpp2015/pdfs/eixo4/a-pobreza-e-a-riqueza-na-regiao-amazonica-e-a-contribuicao-da-politica-de-assistencia-social-o-estado-do-amazonas-em-foco.pdf.

